# Interpretable MRI-Based Deep Learning for Alzheimer’s Risk and Progression

**DOI:** 10.1101/2025.05.06.25326606

**Authors:** Bin Lu, Yan-Rong Chen, Rui-Xian Li, Ming-Kai Zhang, Shao-Zhen Yan, Guan-Qun Chen, Francisco Xavier Castellanos, Paul M. Thompson, Jie Lu, Ying Han, Chao-Gan Yan

## Abstract

Timely intervention for Alzheimer’s disease (AD) requires early detection. The development of immunotherapies targeting amyloid-beta and tau underscores the need for accessible, time-efficient biomarkers for early diagnosis. Here, we directly applied our previously developed MRI-based deep learning model for AD to the large Chinese SILCODE cohort (722 participants, 1,105 brain MRI scans). The model — initially trained on North American data — demonstrated robust cross-ethnic generalization, without any retraining or fine-tuning, achieving an AUC of 91.3% in AD classification with a sensitivity of 95.2%. It successfully identified 86.7% of individuals at risk of AD progression more than 5 years in advance. Individuals identified as high-risk exhibited significantly shorter median progression times. By integrating an interpretable deep learning brain risk map approach, we identified AD brain subtypes, including an MCI subtype associated with rapid cognitive decline. The model’s risk scores showed significant correlations with cognitive measures and plasma biomarkers, such as tau proteins and neurofilament light chain (NfL). These findings underscore the exceptional generalizability and clinical utility of MRI-based deep learning models, especially in large and diverse populations, offering valuable tools for early therapeutic intervention. The model has been made open-source and deployed to a free online website for AD risk prediction, to assist in early screening and intervention.

## INTRODUCTION

Alzheimer’s disease (AD) is a progressive neurodegenerative disease that poses a significant challenge to global health, profoundly impacting individuals, families, and healthcare systems^1^. Early detection of AD is crucial, as it allows for timely interventions that could slow disease progression and improve patient outcomes^1^. The US Food & Drug Administration recently granted traditional approval for lecanemab and donanemab for treating mild cognitive impairment (MCI) and mild AD in patients with amyloid-beta (Aβ) proteinopathy or pathologic tau deposition^2,3^. The advent of these novel immunotherapies highlights the need for cost-effective and time-efficient biomarkers for early diagnosis to enhance treatment effectiveness^4^. Traditionally, AD diagnosis has relied on observing cognitive decline, which typically occurs in later stages, often missing the critical window for effective intervention. Since biological changes often precede cognitive symptoms, integrating biomarkers from cerebrospinal fluid (CSF) and positron emission tomography (PET) into AD diagnostic guidelines has significantly enhanced early detection^5^. However, the invasive nature, high costs and need for specially trained personnel to implement these methods restrict their widespread clinical application.

Alternatively, magnetic resonance imaging (MRI) could provide a non-invasive and more accessible substitute. Currently, structural brain MRI is primarily used to monitor the dynamic progression of AD pathology, including staging, predicting prognosis, and patient selection and safety monitoring during drug treatment^6,7^. However, the advent of deep learning methods has revolutionized MRI’s potential by enabling the detection and use of subtle features previously unrecognizable by human experts, thus advancing the field towards earlier and more precise diagnostics and facilitating timely medical treatment^8–10^. The development of interpretable models has opened the “black box” of deep learning, further enhancing the confidence of physicians and patients in medical imaging models applied to clinical practice^11–13^.

Despite enthusiasm for deep learning in AD diagnosis, significant limitations exist. First, robust independent validation is often lacking. Recent systematic reviews of MRI-based AD classifiers revealed startling statistics: only 0–5.3% of studies (e.g., 2 of 81^14^; 0 of 16^15^; and 6 of 114^16^) used any independent validation dataset, never mind including cross-ethnic validation.

Second, many classifiers have been trained on relatively small sample sizes, leading to models that are susceptible to overfitting. These two factors have contributed to poor generalizability, rendering many models impractical for clinical application. In response to these challenges, we employed transfer learning to construct a generalizable AD classification model based on large and diverse structural MRI datasets, encompassing over 85,000 samples from 217 different sites/scanners^17^. This model achieved an accuracy rate exceeding 90% across four independent AD datasets. Nevertheless, the prior study identified several areas requiring further work: first, the training and validation datasets primarily comprised Western samples, raising concerns about generalizability to other populations, such as those in China; second, the model’s capability to predict disease progression needed further development, as this is essential for selecting patients with early-stage AD for timely pharmacological treatment; third, the pathological relevance of an imaging-based model should be validated by comparison with established biomarkers and cognitive measurements; finally, providing individualized AD-risk brain maps can enhance the model’s interpretability and facilitate monitoring treatment efficacy. Recently, plasma-based biomarkers have shown both feasibility and high accuracy in detecting AD pathology and predicting progression of cognitive decline^18,19^. In the 2024 revision of the NIA-AA diagnostic criteria, plasma analytes such as phosphorylated tau 217 immunoassay were designated as belonging to the “core-1” category, equivalent to CSF and Amyloid PET^20^. Cross-validating these two types of clinical feasibility biomarkers (plasma-based and MRI-based metrics) holds significant practical implications.

To achieve these objectives, we applied an MRI-based classification model to a non-Western, longitudinal, richly-phenotyped cohort, SILCODE, which consists of a large elderly Chinese population, assessed across the stages of the AD continuum^21^. This cohort includes extensive cognitive assessments, plasma biomarkers, and brain images. In the current study, we validated the MRI-based AD model based on its classification accuracy, predicting patients’ disease progression, and its association with cognitive and pathological markers using the SILCODE sample. Additionally, we developed an interpretable method to enhance its clinical credibility.

## MATERIALS AND METHODS

### Participants and procedures

We included participants from the Sino Longitudinal Study on Cognitive Decline (SILCODE), a registered ongoing longitudinal clinical trial (ClinicalTrials.gov identifier: NCT02225964) focused on AD in the elderly Han population in mainland China. This study was approved by the ethics committee of Xuanwu Hospital of Capital Medical University, and all participants or their guardians provided written informed consent before enrollment. The detailed SILCODE protocol has been previously described^22^. Participants were continuously recruited and followed up with flexible visit intervals, initially set at 15 months and adjusted based on cognitive evaluations administered by phone at three-month intervals, leading to varying follow-up times and visit intervals for participants. During each visit, participants were invited to Xuanwu Hospital for clinical evaluation, neuropsychological assessments, blood tests, and MRI scans.

The current study initially included 738 participants, all over age 50 years at the time of their first MRI scan, who had completed at least one MRI session. After quality control, 16 participants were excluded due to invalid MRI scans, leaving a final sample of 722 participants (60.8% female; mean age = 66.1 ± 7.7 years at baseline). Diagnostic distribution at baseline was: 211 normal cognition (NC), 235 subjective cognitive decline (SCD), 193 MCI, and 83 AD. Participants and their visit sessions were further filtered based on the requirements of specific analyses.

### Diagnostic criteria

During each visit, participants were classified into AD, MCI, SCD, and NC groups, with re-evaluations in subsequent visits to detect changes^22^. AD diagnosis followed NIA-AA guidelines^23^, supplemented by a global Clinical Dementia Rating^24^ (CDR) score of at least 1.

MCI was diagnosed using the Jak/Bondi method^25^ with neuropsychological tests (see below) and thresholds adjusted for the Chinese population^22^. SCD classification followed Jessen et al.’s criteria^26^ for SCD plus. NC was assigned to those without subjective or informant-reported memory decline or evidence of cognitive impairment. In overlapping cases, the more severe condition was designated. All clinical diagnoses were made by qualified physicians.

### Neuropsychological tests

A battery of neuropsychological tests was used to measure cognitive levels of participants, focusing on both global domain and specific subdomains including memory, language, and executive functions. Global cognitive function was assessed using the Mini-Mental State Examination^27^ (MMSE), Memory and Executive Screening^28^ (MES), and the Montreal Cognitive Assessment Basic^29^ (MOCA-B). For specific subdomains, memory tests included the Auditory Verbal Learning Test—Huashan version^30^—Long delayed memory (AVLT-L) and Recognition (AVLT-R); the language tests comprised the Animal Fluency Test^31^ (AFT) and 30-item Boston Naming Test^32^ (BNT); and executive functions were evaluated using the Shape Trail Test^33^ Parts A (STT-A) and B (STT-B). All cognitive assessments were administered by trained neuropsychologists.

### Biological tests

Plasma levels of Aβ42, Aβ40, NfL, GFAP, p-tau 217, and p-tau 181 were measured by immunoassay using a Simoa HD-X analyzer (Quanterix, Billerica, MA, USA), as per the manufacturer’s protocols. Specifically, plasma samples were thawed at room temperature and then centrifuged for 5 minutes at 4°C and 10,000 × g. The resultant supernatant was transferred to 96-well plates and diluted four-fold with sample diluent, followed by a two-step digital immunoassay. The concentrations of Aβ42, Aβ40, NfL, and GFAP were measured with the Simoa Neurology 4-Plex E Advantage Kit (Quanterix, Billerica, MA, USA; Cat# 103670). The ALZpath Simoa pTau-217 v2 Assay Kit (Quanterix, Cat# 104371) was used to measure p-tau 217, and the Simoa pTau-181 Advantage V2 Kit (Quanterix, Cat# 103714) was used to measure p-tau 181. Assays were performed using kits from the same lot, with all samples run in duplicate. Seven or eight calibrators (2x) and two quality controls (2x) were run on each plate for each analyte. The lower limits of detection were 0.136 pg/mL for Aβ42, 0.384 pg/mL for Aβ40, 0.090 pg/mL for NfL, 0.441 pg/mL for GFAP, 0.019 pg/mL for p-tau 217, and 0.028 pg/mL for p-tau 181. The lower limits of quantification were 0.378 pg/mL for Aβ42, 1.02 pg/mL for Aβ40, 0.400 pg/mL for NfL, 2.89 pg/mL for GFAP, 0.061 pg/mL for p-tau 217, and 0.338 pg/mL for p-tau 181. The intra-assay coefficients of variation for controls ranged from 2% to 13% for Aβ42, 1% to 5% for Aβ40, 2% to 12% for NfL, 1% to 8% for GFAP, 2% to 11% for p-tau 217, and 1% to 10% for p-tau 181.

### MRI acquisition, preprocessing, and AD risk score generation

T1-weighted structural brain MRI data were collected using a single 3.0 T GE SIGNA^TM^ PET/MR scanner (GE Healthcare, Milwaukee, WI, USA) at Xuanwu Hospital of Capital Medical University. Scanning parameters were as follows: SPGR sequence with inversion preparation, repetition time (TR)=6.9 ms, echo time (TE)=2.98 ms, inversion time (TI)=450 ms, slice number=192, slice thickness=1 mm, gap=0, flip angle=12°, field of view (FOV)=256×256 mm^2^, matrix=256×256, voxel size=1×1×1 mm^3^.

Manual quality control was conducted to exclude low-quality images with notable artifacts or incomplete brain coverage. Images were then preprocessed using the sMRI preprocessing pipeline from the Data Processing Assistant for Resting-State fMRI (DPARSF) Toolbox. GMD and GMV maps were generated using the Voxel-Based Morphometry (VBM) module in the toolbox. These maps were then directly input to the deep learning AD classifier to compute AD risk scores. More information on preprocessing, feature extraction, and risk score generation processes is available in the previous study^34^ on the AD classifier.

In this study, two types of risk scores were computed: sigmoid and linear. Sigmoid risk scores were obtained using sigmoid activation in the deep learning model, as per the previous AD classifier paper^34^. Linear risk scores, produced using linear activation, were additionally calculated due to their distribution being more suitable for statistical testing, in that they are closer to the normal distribution than sigmoid scores. Accordingly, we used sigmoid scores for classification-related analysis, and linear scores for statistical tests such as linear regression.

### Data analysis

We first evaluated the performance of the MRI-based AD classifier on the Chinese SILCODE cohort using receiver operating characteristic (ROC) curve analysis. The area under the curve (AUC) was calculated, with 95% confidence intervals (CIs) estimated using 5,000 bootstrap samples. The optimal cutoff score was determined using the Youden index^35^. At this threshold, we calculated accuracy, sensitivity and specificity to assess classifier performance.

Next, we applied a linear mixed-effects model to examine the association between classifier linear scores and clinical diagnoses (NC, SCD, MCI, AD), controlling for sex, age, and education. Participants were included as a random effect to account for the non-independence of repeated measures, as some participants had multiple visits. Post-hoc pairwise comparisons with Tukey correction were performed to examine differences between diagnostic groups. The Satterthwaite approximation was used to adjust degrees of freedom for fixed effects, accounting for the uncertainty introduced by random factors, to provide more robust results. Post-hoc pairwise comparisons were conducted using estimated marginal means with Tukey correction to examine differences between diagnostic groups.

Survival analyses were conducted to examine the probability of progression to AD in NC, SCD and MCI participants. Participants were divided into high-risk and low-risk groups based on AD classifier risk scores at baseline, with the cutoff (i.e., 0.38) identified through ROC analysis using the Youden index. Kaplan-Meier survival curves were generated to illustrate the probability of participants maintaining non-AD status over the follow-up period, with the log-rank test comparing survival distributions between groups.

Additionally, visit sessions from all AD converters were analyzed to assess how early the classifier could predict conversion. The proportion of high-risk sessions (score ≥ 0.38) at different time intervals before the first AD diagnosis was used to calculate the capture rate. Mean risk scores were also computed to observe trends in AD risk over time. Due to the limited data, particularly the decrease in data size as the time before diagnosis increased, we categorized time intervals to simplify analyses. Intervals were set as within 1 year, 1–2 years, 2–3 years, 3–5 years, and 5–14 years before the first AD diagnosis (**Figure S1**).

To examine the associations of MRI-based risk scores with cognitive tests and plasma biomarkers, separate models were fitted for each measure. Cognitive and plasma outliers exceeding six times the interquartile range (6IQR)^36^ were excluded to minimize their influence. Global associations were first assessed across the entire sample to identify general patterns, followed by diagnosis-specific analyses within pre-AD phases (NC, SCD, MCI) to explore phase-specific associations critical for early detection and intervention. Linear mixed-effects models were generally used, with the linear risk score as the independent variable, sex, age, and education as covariates, and participant included as a random effect to account for repeated measures. However, for plasma biomarker models within the MCI group, general linear models without the random effect were applied due to the limited number of repeated measures per participant (e.g., 34 participants had only 37 plasma p-tau 217 measures), which caused convergence issues in mixed-effects models. Effect sizes were measured using Cohen’s *f^2^*, which reflects the extent to which the inclusion of a specific predictor variable improves the model’s explanatory power. The Satterthwaite approximation was used to adjust degrees of freedom for fixed effects in mixed-effects models. Post-hoc pairwise comparisons were performed using the estimated marginal means with Tukey correction in all models.

Differences across brain subtypes (see the next section for details on identification) in variables of interest, such as MMSE decline rate, baseline MMSE level, baseline linear risk score, and demographic variables, were assessed using ANOVA, followed by post-hoc pairwise comparisons of estimated marginal means with LSD correction.

All analyses and visualizations were conducted using R 4.4.0^37^, along with the following packages: pROC^38^ (for ROC analysis), lme4^39^ and lmerTest^40^ (for linear mixed models), survival^41^ and survminer^42^ (for survival analysis), and emmeans^43^ (for post-hoc comparisons).

### Identifying high-risk brain regions associated with AD

We developed an interpretable approach to identify brain regions at risk for AD on an individual level. An AD brain risk map can be generated for each NC, SCD, MCI, or AD participant, in which brain regions with higher values are more closely associated with AD. The detailed calculation process is as follows.

Initially, we created a simulated patient whom the classifier identified as being in an exact intermediate state between AD and NC. To achieve this goal, we conducted 10,000 random samplings within the AD and NC samples of the ADNI 1, ADNI GO and ADNI 2 datasets^44^. For each sampling, we randomly selected 100 samples (e.g., 48 samples from the AD group and 52 samples from the NC group) and averaged their GMD and GMV maps to generate an average template. Subsequently, the MRI-based AD classifier^34^ was used to predict AD risk scores for 10,000 simulated patients. The GMD and GMV maps of the simulated patient whose risk score was closest to 0.5 were selected as the final brain risk map template (the AD risk score for the final template was 0.499928).

To predict the AD risk regions for a single participant, we sequentially replaced region by region in the aforementioned simulated template with the corresponding real data from the participant’s brain. Then, we used the MRI-based AD classifier to predict the AD risk score of the reconstituted brain. For instance, when the left hippocampal-related brain region of an AD patient was replaced onto the simulated template, the predicted AD risk score rose from 0.5 to 0.88, indicating that the left hippocampus’s area risk level for that patient was 0.88. Repeating this process and then reprojecting the risk scores of every brain area back onto the brain generates the subject’s AD risk distribution map. Each replaced brain region was defined as a cube with an edge length of 18 mm, and the step length for moving and replacing brain regions was 12 mm.

Subsequently, we employed k-means cluster analysis to examine different AD risk brain region patterns within the AD patients in the SILCODE dataset. We determined the optimal number of clusters for k-means clustering by employing the elbow method, which utilizes the sum of squared errors (SSE) to identify the point at which the increase in the number of clusters does not result in significantly better modeling of the data. The center of the feature space for each cluster is considered to be a representative pattern of AD-related brain abnormalities, or alternatively, a demonstration of an AD subtype based on MRI characteristics.

## RESULTS

### Study participants

The current study included 722 participants from the SILCODE dataset (60.8% female; 66.1 ± 7.7 years of age at baseline), categorized based on clinical assessments at baseline into 211 NC, 235 SCD, 193 MCI, and 83 AD participants. They completed 1,980 visits and provided 1,105 valid MRI scans. **Table 1** presents descriptive data for these MRI sessions (60.8% female; 66.6 ± 7.6 years; 348 NC, 363 SCD, 269 MCI, 125 AD), which serve as the basis for the following validations of classificatory ability and pathological value of the MRI-based deep learning model.

**Table 1.**
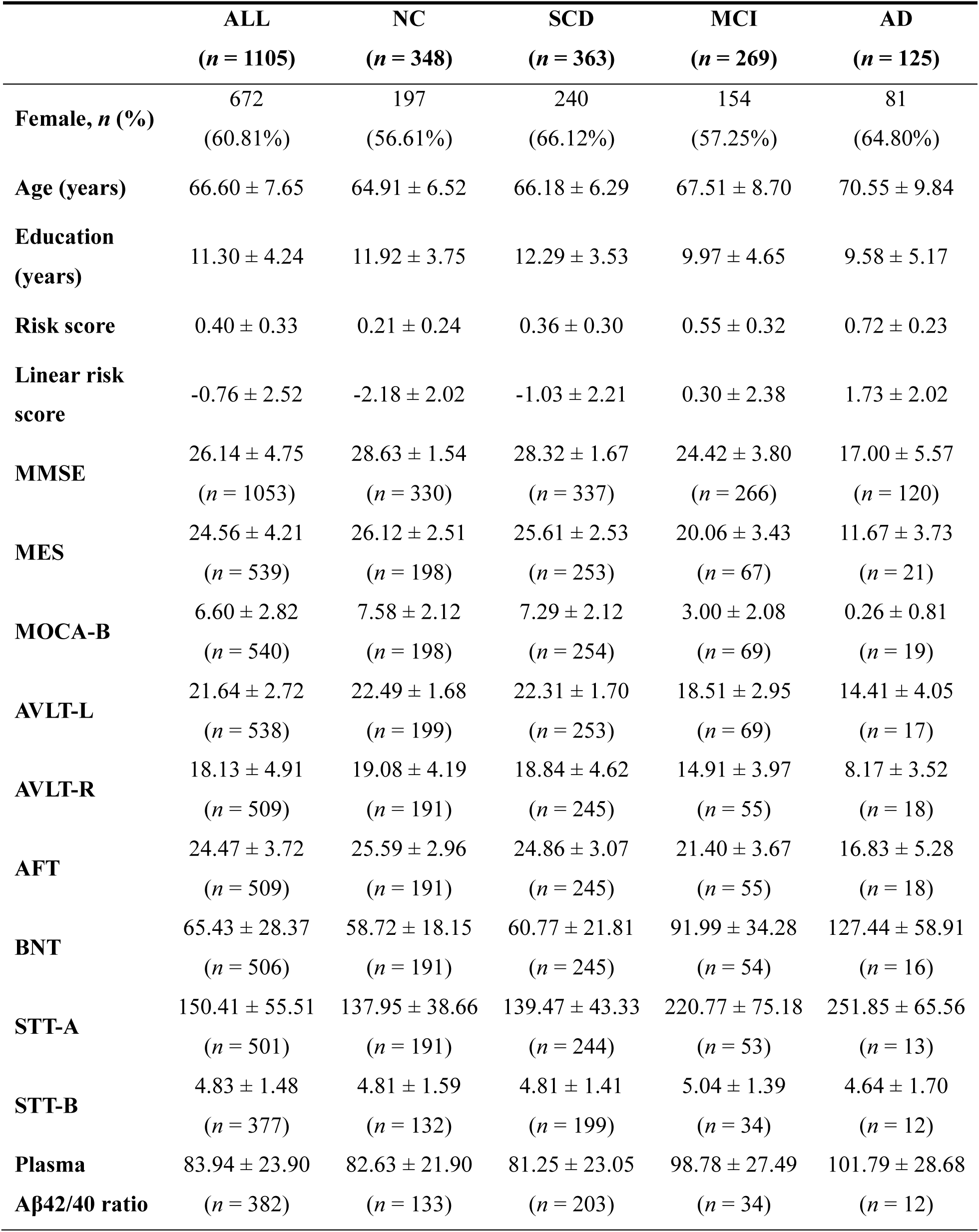
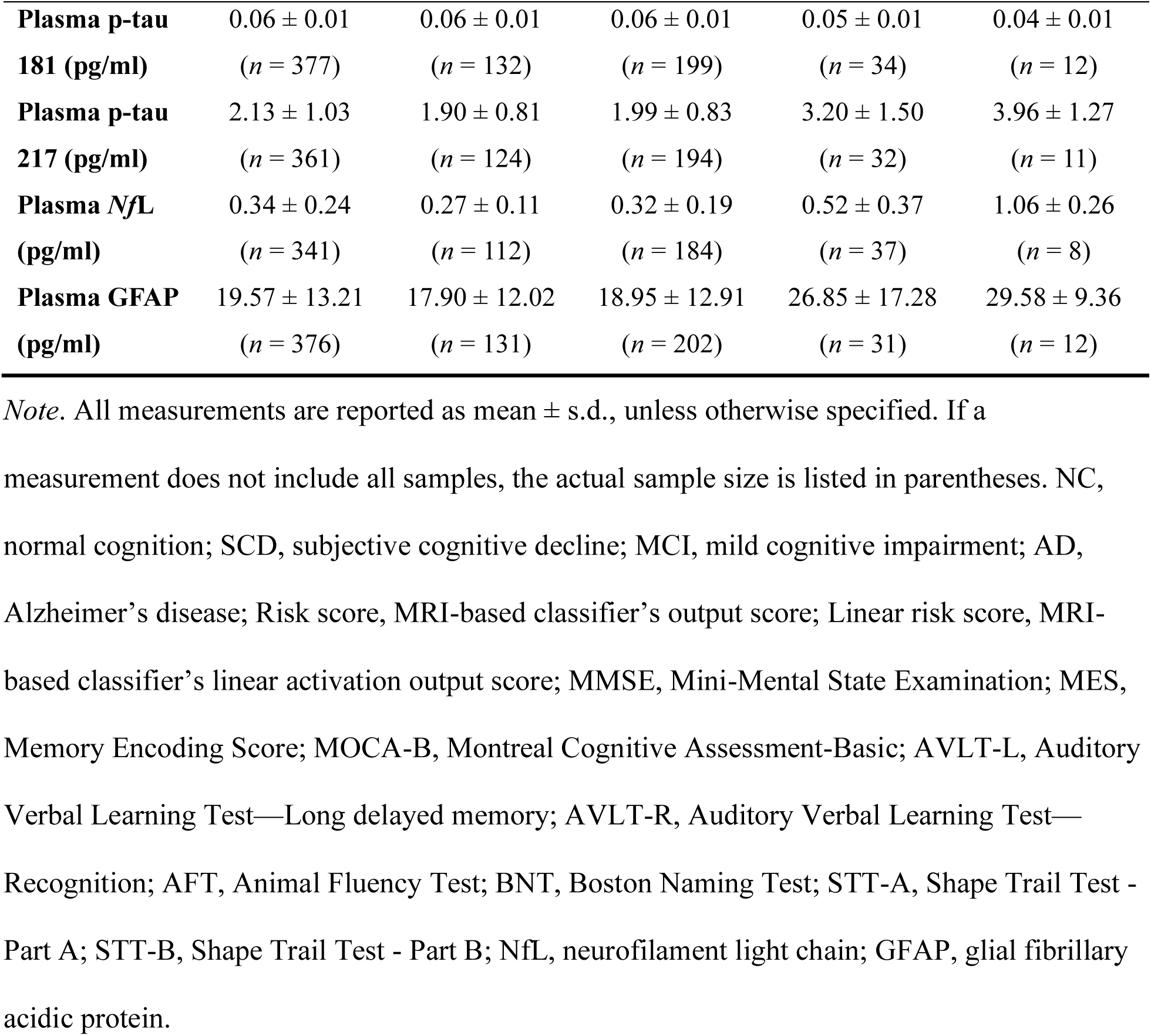
Descriptive Data for Visit Sessions with Valid MRI Scans.

### AD classification performance in a Chinese sample

When directly applied to the SILCODE cohort without any re-training or fine-tuning, the MRI-based AD classifier demonstrated robust performance in distinguishing AD from NC. As **Fig. 1a** shows, the area under the curve (AUC) was 91.3% (95% CI = [88.3%, 94.0%]), indicating solid discriminative capability. An optimal cutoff score of 0.38 was identified using the Youden index. At this threshold, the classifier achieved an accuracy of 88.2%, a sensitivity of 95.2%, and a specificity of 85.6%.

**Fig. 1.**
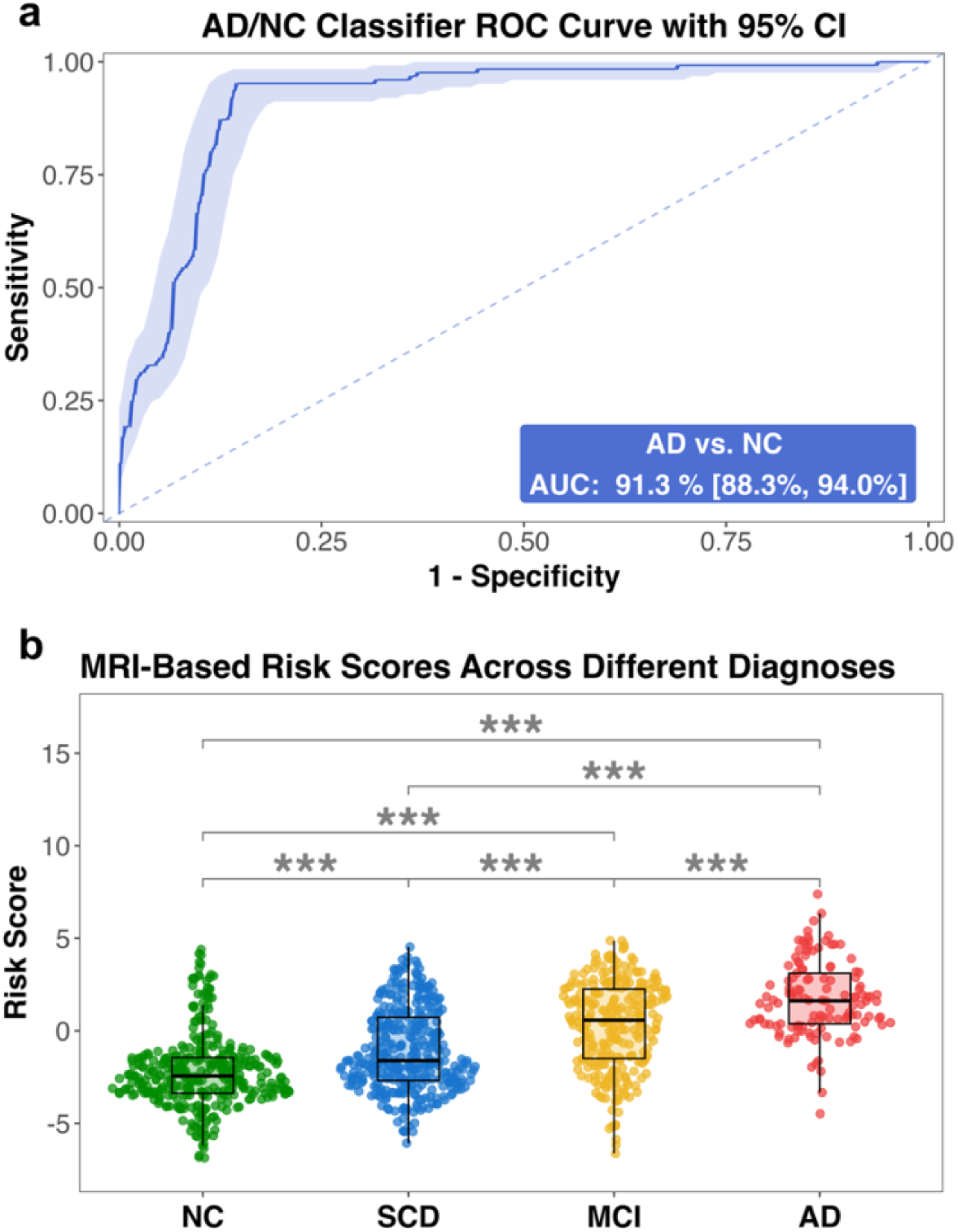
Classification performance of the MRI-based AD classifier on the SILCODE Cohort. *Note*. **a,b,** Performance of of the MRI-based AD classifier. (a) The receiver operating characteristic (ROC) curve shows the classifier’s performance in distinguishing Alzheimer’s disease (AD) from normal cognition (NC), with an area under the curve (AUC) of 91.3% (95% confidence interval [CI]: 88.3%, 94.0%). The shaded area represents the 95% CI. (b) Box plots show the median, interquartile range, and whiskers extending 1.5 times the interquartile range, with individual data points displayed. \*\*\**p* < 0.001.

### Classifier scores correspond to clinical diagnoses

Adjusting for sex, age, and education, and accounting for non-independent samples by including participant as a random effect, we found that linear risk scores^1^ varied significantly by clinical diagnoses (*F*(3, 1058.04) = 73.96, *p* < 2.20 × 10^-16^). Post-hoc Tukey-corrected comparisons revealed that the AD group had significantly higher scores than NC (mean difference = 3.36, *SE* = 0.24, *t*(1018.86) = 14.11, *p* = 7.85 × 10^-13^), SCD (mean difference = 2.30, *SE* = 0.23, *t*(1045.43) = 9.84, *p* < 2.20 × 10^-16^), and MCI groups (mean difference = 1.23, *SE* = 0.22, *t*(1051.45) = 5.61, *p* = 1.57 × 10^-7^). MCI also scored higher than NC (mean difference = 2.13, *SE* = 0.19, *t*(947.90) = 11.10, *p* = 3.35 × 10^-13^) and SCD (mean difference = 1.07, *SE* = 0.19, *t*(1036.81) = 5.78, *p* = 6.05× 10^-8^). In addition, SCD scored higher than NC (mean difference = 1.06, *SE* = 0.17, *t*(1038.16) = 6.78, *p* = 6.05× 10^-8^). As shown in **Fig. 1b**, the relative ordering of scores across diagnostic groups aligned with expected severity along the AD continuum, despite the classifier not being trained or fine-tuned on SCD or MCI data.

### Performance in predicting disease progression

A total of 474 participants, comprising 124 with MCI, 195 with SCD, and 155 with NC as diagnosed at their baseline visits, who completed at least two visit sessions, were included in the survival analysis. Participants were further classified as high-risk (risk score ≥ 0.38) or low-risk (risk score < 0.38) based on their baseline sigmoid risk scores. In the full sample, 60 participants converted to AD during the 167-month follow-up period, including 57 conversions from the baseline MCI group and 3 from the baseline SCD group. No baseline NC participant converted to AD during the tracking period. Kaplan-Meier survival plots illustrate the detailed progression of AD conversion in the full sample (**Fig. 2a**), baseline MCI group (**Fig. 2b**), and baseline SCD group (**Fig. 2c**). Notably, as shown in the table at the bottom of **Fig. 2a**, our model identified 52 of 60 AD converters as high risk at baseline, achieving an overall capture rate of 86.7%. In the full sample, 52 of the 177 participants classified as high-risk and 8 of the 297 participants classified as low-risk converted to AD during the follow-up period. The log-rank test indicated a significant difference between the survival curves (*p* < 0.0001), accounted for by higher probability of conversion for participants in the high-risk group.

**Fig. 2.**
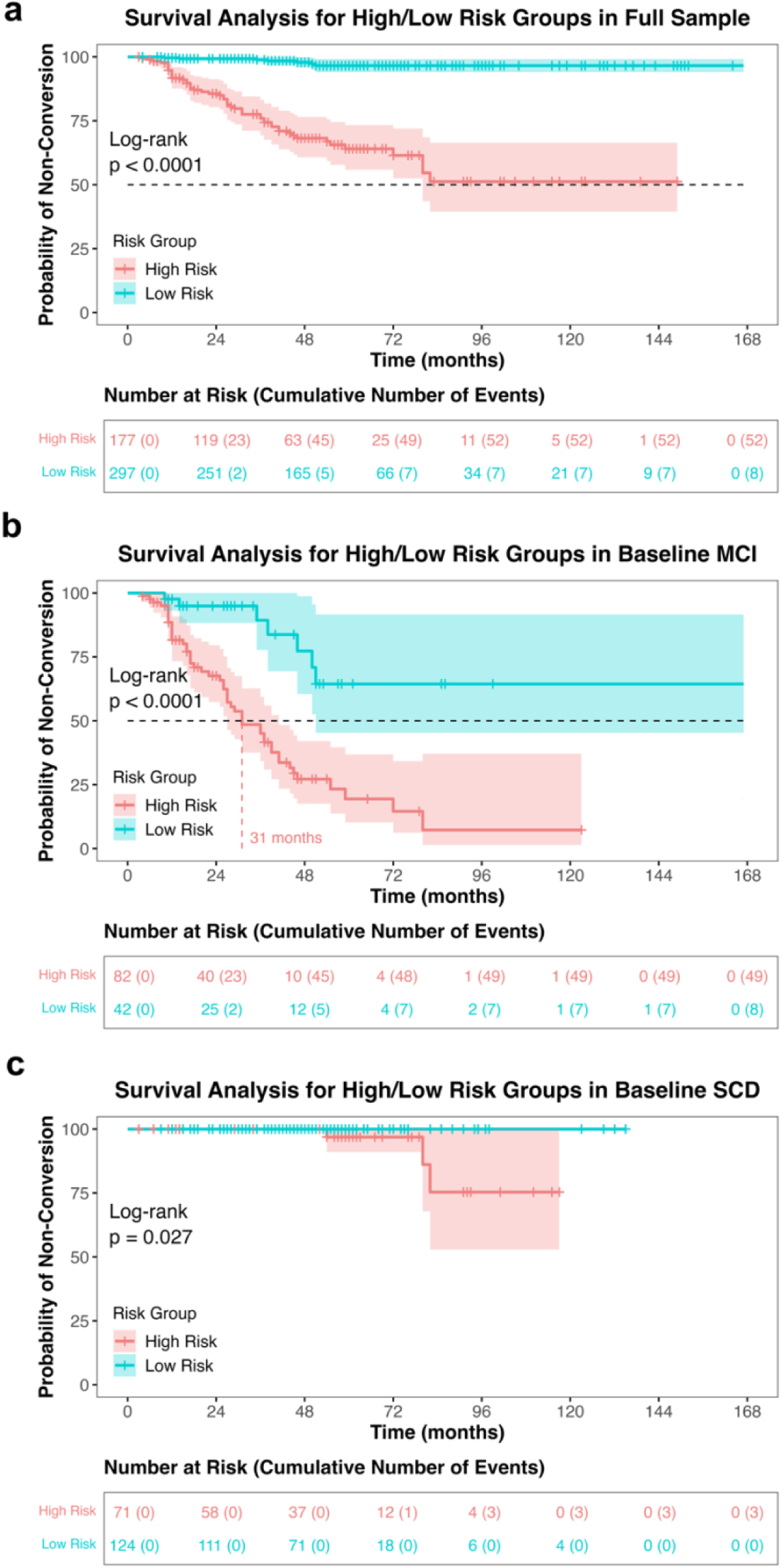
Survival analysis for baseline MCI and SCD groups. *Note*. **a, b, c,** Kaplan-Meier survival (conversion) curves showing the probability of non-conversion in high-risk (red) and low-risk (blue) groups over 167 months (the longest follow-up period). High risk was defined by baseline AD sigmoid risk scores (≥0.38). Curves are shown for the full sample (**a**), MCI (**b**), and SCD (**c**) at baseline. The baseline NC sample is not shown, as no baseline NC participant progressed to AD during the tracking period. Shaded areas represent 95% confidence intervals, with vertical ticks indicating censored participants (lost to follow-up). Vertical dotted lines mark median conversion times, and the horizontal black dashed line indicates a 50% probability of non-conversion. Log-rank tests compared survival (conversion) distributions between the two groups, with *p* < 0.05 indicating significance. Tables below the plots show the number of participants at risk (i.e., still followed and non-progressed) at each time point, with cumulative conversion events in parentheses. To avoid confusion, the survival curves for the low-risk group (**a** and **b**) omit the abrupt drop to zero at the tail caused by the last single individual’s conversion to AD. MCI, mild cognitive impairment; SCD, subjective cognitive decline.

Specifically, among participants with MCI at baseline (**Fig. 2b**), 49 of the 82 high-risk participants converted to AD, compared to 8 of the 42 low-risk participants, showing a significantly higher probability of conversion to AD in the high-risk group (log-rank test, *p* < 0.0001). The median survival time (i.e., time-to-conversion) of the high-risk MCI group (31 months) was significantly shorter than that of the low-risk MCI group (>167 months). In participants with SCD at baseline, only 3 conversions occurred, and all were in the high-risk category (**Fig. 3c**). The log-rank test indicated a significantly higher risk of conversion for high-risk SCD participants (*p* = 0.027, **Fig. 2**). In summary, survival analyses demonstrated that participants identified as high-risk at baseline by our classifier were more likely to convert to AD over time.

**Fig. 3.**
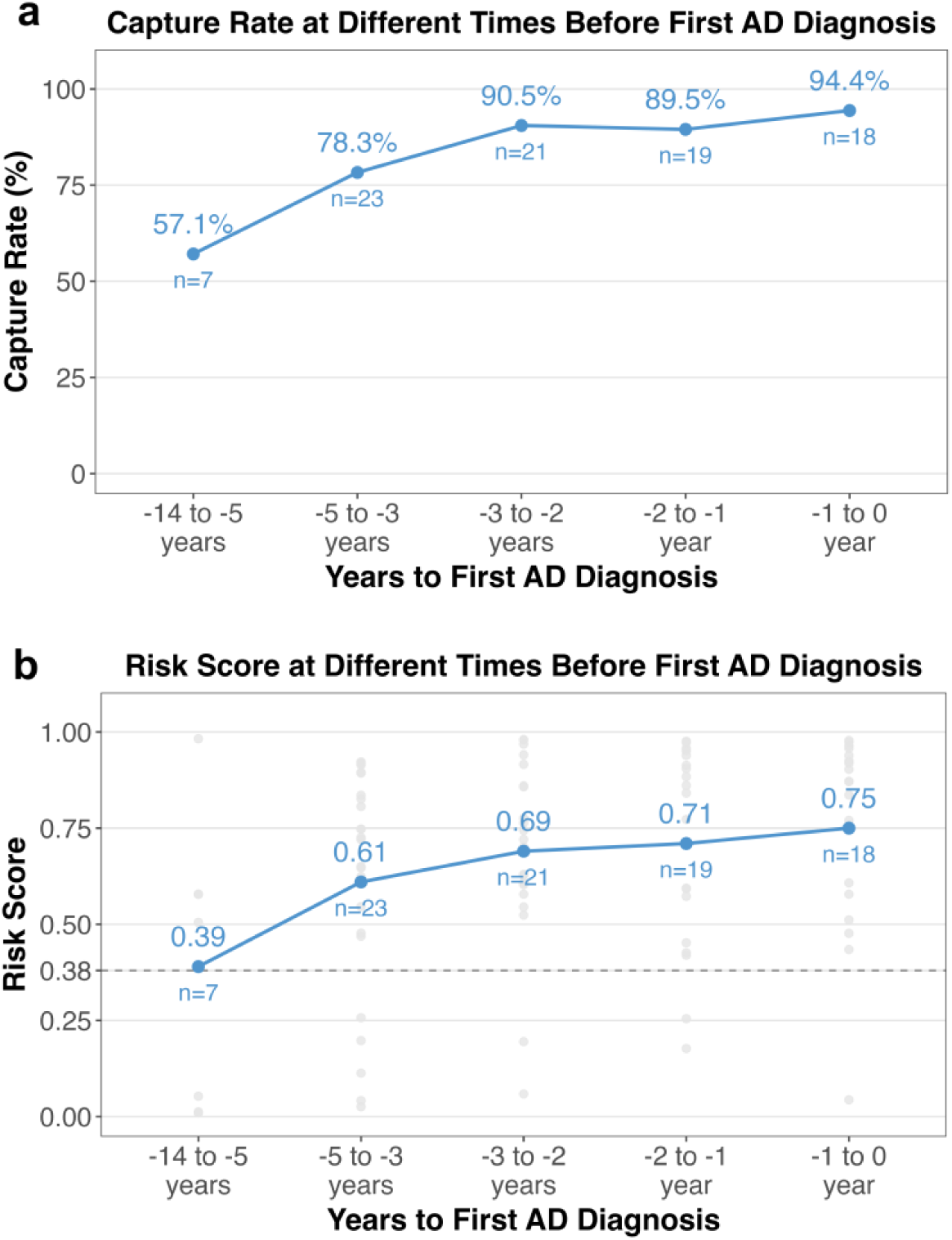
Proportion exceeding high-risk criteria and mean sigmoid risk scores before AD diagnosis. *Note.* **a**, **b**, The proportion of sessions with sigmoid risk scores meeting the high-risk threshold (≥ 0.38) (**a**) and the mean sigmoid risk scores (**b**) across different time intervals before the first AD diagnosis. The numbers in **a** represent the proportions, while the numbers in **b** indicate the mean risk scores. *n* refers to the number of available sessions in each time period. The horizontal dotted grey line in **b** marks the 0.38 cut-off for high risk. AD, Alzheimer’s disease.

### Classifier predicted AD progression years before diagnosis

Using all visit sessions (*n* = 88) from the 60 participants who progressed from NC, SCD, or MCI, we estimated *how early* the classifier was able to predict AD conversion. Specifically, we calculated the ratio of sessions flagged as high risk by the classifier (risk score ≥ 0.38) at various time intervals before the first AD diagnosis. Results (**Fig. 3a**) show that the classifier achieved a capture rate of about 90% within 3 years, 78% within 3–5 years, and 57% within 5–14 years before the first AD diagnosis among AD converters. Mean risk scores increased monotonically across follow-up periods, consistent with the probability of conversion (**Fig. 3b**).

### Risk scores associated with cognitive assessments and plasma biomarkers

Linear mixed-effects models were employed to examine the associations of MRI-based AD risk scores with standardized cognitive assessments and plasma biomarker levels. Global associations were first assessed across all samples, without distinguishing diagnostic phases, to identify general patterns. Diagnosis-specific analyses were then conducted within each pre-AD diagnostic phase to evaluate more granular associations, given the importance of these phases for understanding early-stage changes critical for detection and intervention.

As shown in **Fig. 4a** and **Table 2**, MRI-based risk scores were significantly associated with all cognitive measures, including both global cognitive assessments (i.e., MMSE, MES, MOCA-B) and subdomain-specific tests (i.e., AVLT-L, AVLT-R, AFT, BNT, STT-A, STT-B). Higher MRI-based risk scores were linked to poorer cognitive performance across the board.

**Fig. 4.**
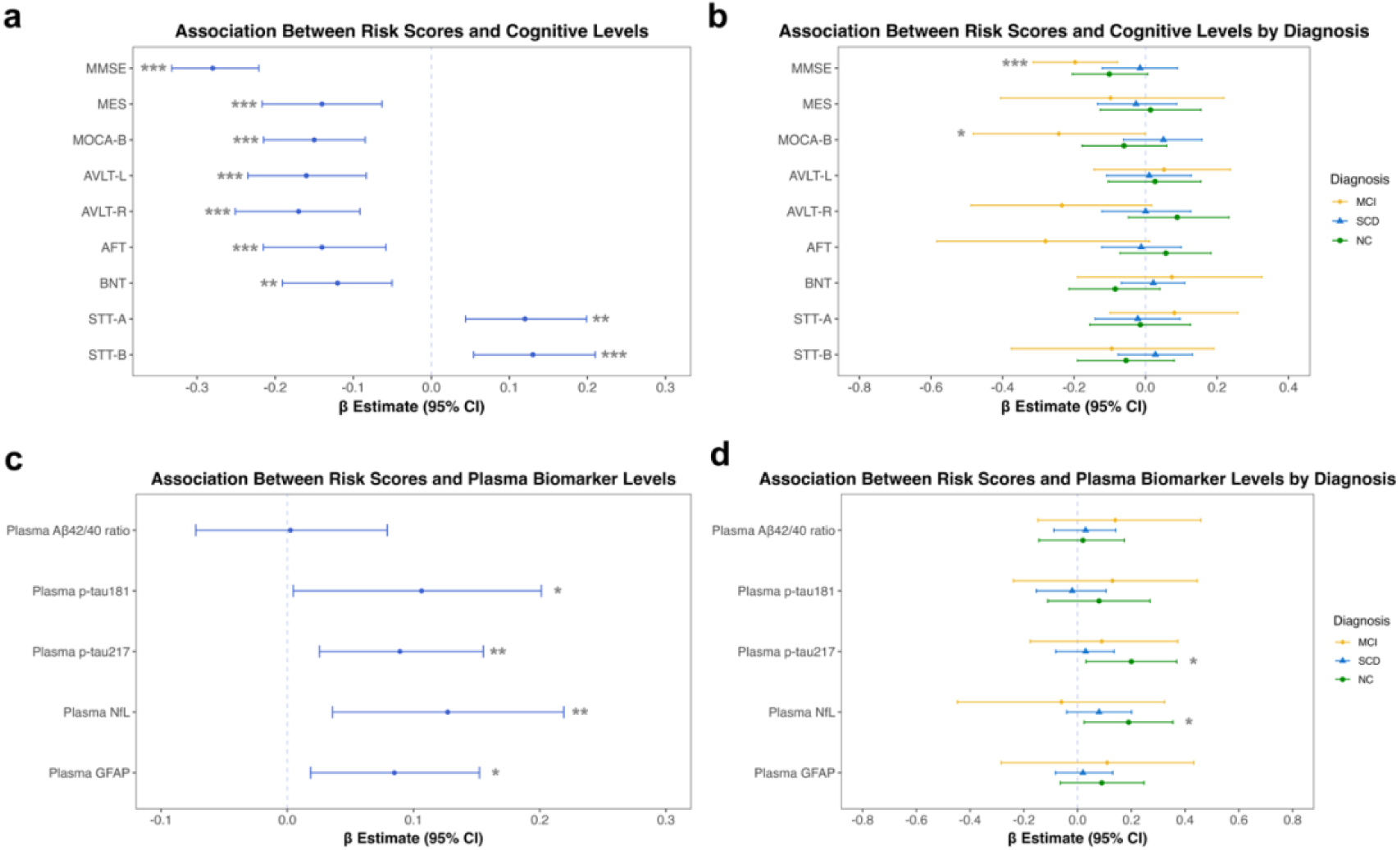
Risk score associations with cognitive performance and plasma biomarkers. **a, b**, Associations between MRI-based risk scores and cognitive performance across all samples (**a**) and within each pre-AD group (**b**). **c, d,** Associations between risk scores and plasma biomarker levels across all samples (**c**) and within each pre-AD group (**d**). Separate models were fitted for each cognitive and plasma measure, with all variables standardized. The linear risk score was the independent variable, while sex, age, and education were included as covariates.

**Table 2.**
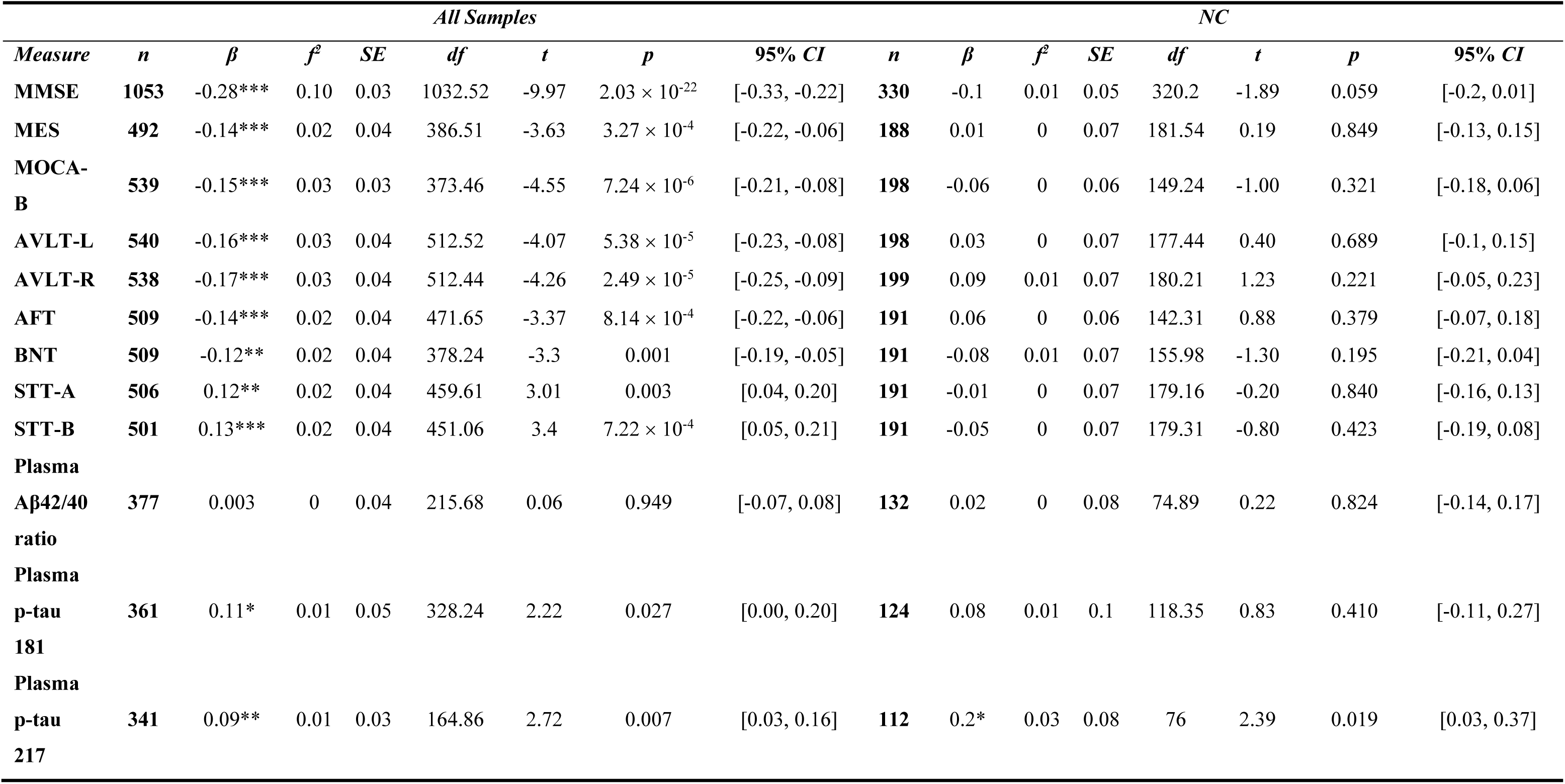
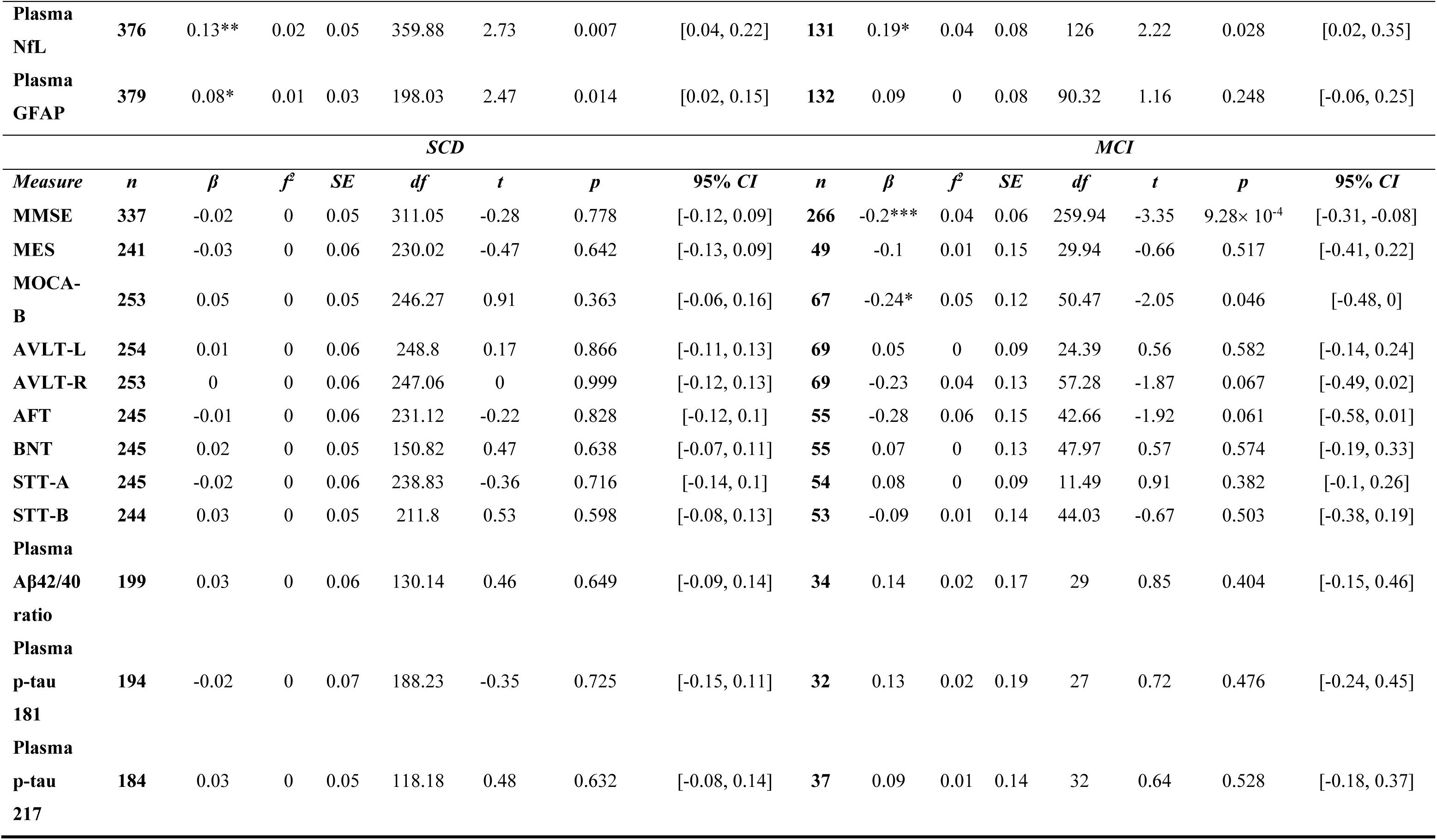
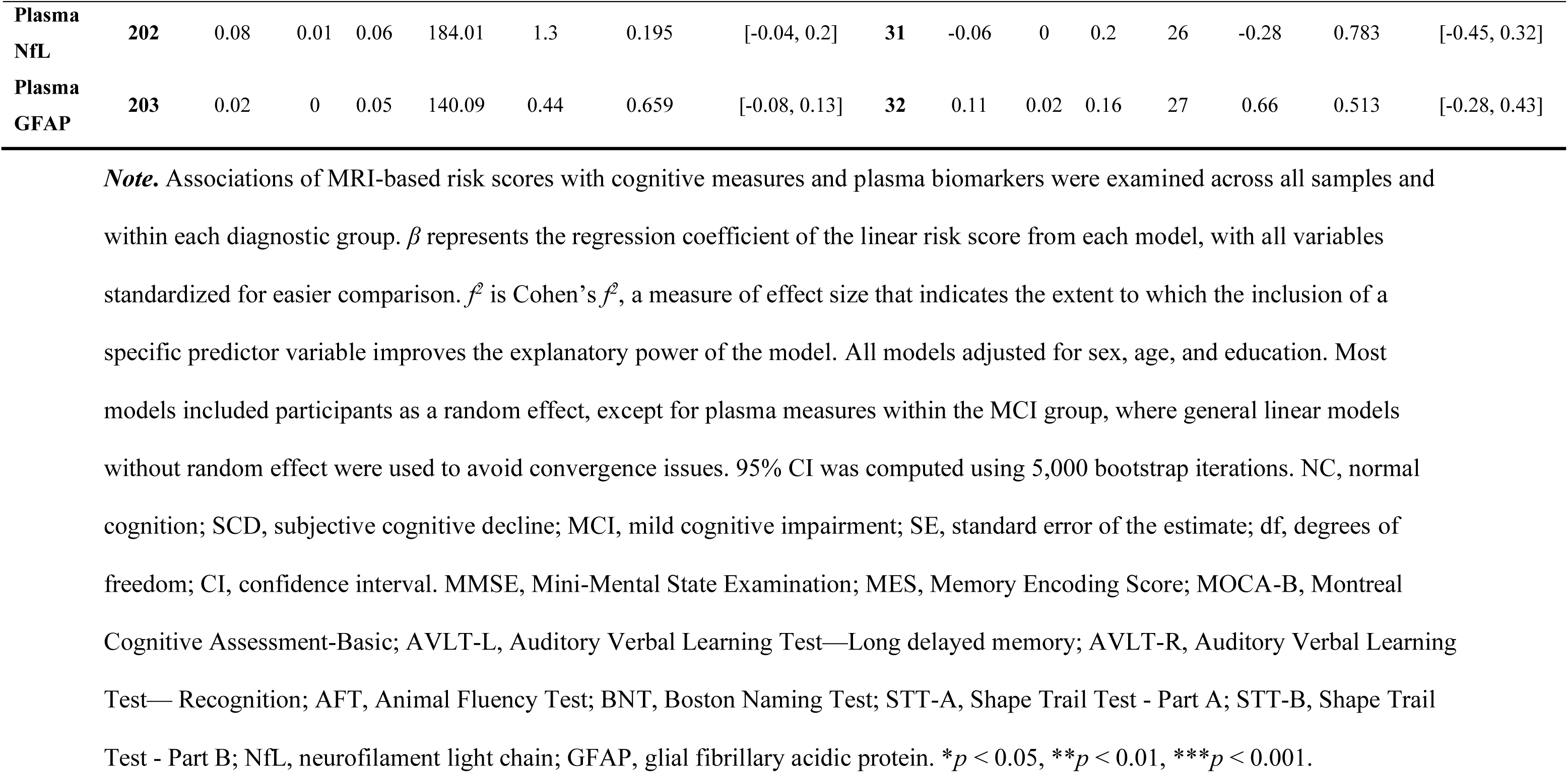
Associations between MRI-Based Risk Scores and Cognitive Measures and Plasma Biomarkers, across All Samples and by Diagnostic Phase.

Specifically, higher scores on the STT-A, and STT-B (where higher scores indicate worse cognitive function) were associated with higher risk scores (*p*s < 0.01, *f^2^*s = 0.02). By contrast, for the remaining cognitive measures, where lower scores indicate worse cognitive function (e.g., MMSE, MES, MOCA-B), higher risk scores were associated with lower scores, indicating a decline in cognitive performance (*p*s < 0.01, *f^2^*s = 0.02 – 0.10). When examined within each diagnostic phase, significant associations between MRI-based risk scores and cognitive performance were observed only within the MCI phase. Specifically, risk scores remained significantly associated with MMSE (*p* < 0.001, *f^2^* = 0.04) and MOCA-B (*p* < 0.05, *f^2^* = 0.05) within the MCI phase.

When plasma biomarkers were examined across all samples (**Fig. 4c** and **Table 2**), higher MRI-based risk scores were positively associated with elevated levels of p-tau 181, p-tau 217, NfL, and GFAP (*p*s < 0.05, *f^2^*s = 0.01 – 0.02), indicating that higher risk scores are linked to increased levels of these neurodegenerative markers. When examined by diagnosis, significant associations between MRI-based risk scores and plasma biomarker levels were observed only within the NC phase. Specifically, risk scores remained significantly associated with p-tau 217 (*p* < 0.05, *f^2^* = 0.03) and NfL (*p* < 0.05, *f^2^* = 0.04). In the SCD and MCI groups, the associations were all non-significant (SCD: *p*s > 0.05, *f*^2^s ≈ 0; MCI: *p*s > 0.05, *f*^2^s = 0–0.02). The associations between risk scores and the plasma Aβ42/40 ratio were non-significant, both across all samples and within diagnoses.

Most models used linear mixed-effects analysis, with participant included as a random effect. However, for plasma measures within the MCI group, general linear models without random effect were used instead due to the limited number of repeated measures per participant, which caused convergence issues in mixed-effects models. Horizontal lines represent 95% confidence intervals (CIs; based on 5,000 bootstrap samples) for the *β* estimates of the linear risk score; CIs that do not include 0 indicate statistical significance. The dotted vertical line represents zero, with estimates to the left indicating negative associations and those to the right indicating positive associations. NC, normal cognition; SCD, subjective cognitive decline; MCI, mild cognitive impairment; AD, Alzheimer’s disease; CI, confidence interval; MMSE, Mini-Mental State Examination; MES, Memory Encoding Score; MOCA-B, Montreal Cognitive Assessment-Basic; AVLT-L, Auditory Verbal Learning Test—Long delayed memory; AVLT-R, Auditory Verbal Learning Test— Recognition; AFT, Animal Fluency Test; BNT, Boston Naming Test; STT-A, Shape Trail Test - Part A; STT-B, Shape Trail Test - Part B; NfL, neurofilament light chain; GFAP, glial fibrillary acidic protein. \**p* < 0.05, \*\**p* < 0.01, \*\*\**p* < 0.001.

### Subtyping and predicting cognitive decline speed via an interpretable model

Individualized AD brain risk maps were generated for each MRI session for all participants. In the AD group, all patients could be clustered into three categories based on the distribution of the risk areas, forming three MRI-based AD subtypes (**Fig. S2**). The essential high-risk brain regions of the three AD subtypes are shown in **Fig. 5a-c**. The primary risk regions for AD subtype 1 were located around the midbrain area, including the midbrain tectum, red nucleus, dorsal raphe nucleus, and pineal gland. The primary risk regions for AD subtype 2 included hippocampus and posterior cingulate cortex. The primary risk regions for AD subtype 3 included hippocampus, the midbrain tectum, red nucleus, dorsal raphe nucleus, and pineal gland. All SILCODE AD patients misclassified as NC by the MRI-based classifier belong to AD subtype 1 (**Fig. 5d**).

**Fig. 5.**
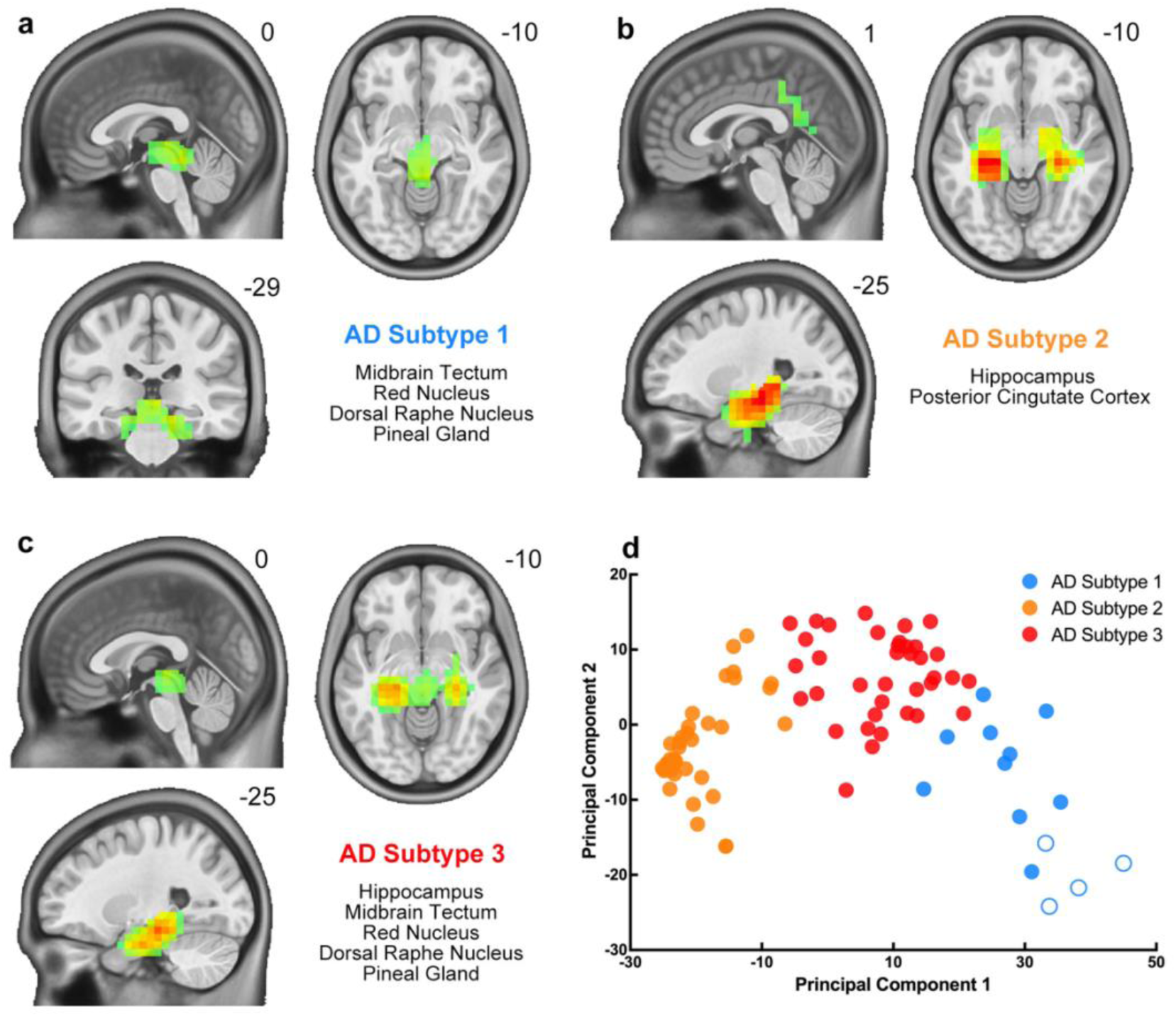
Brain risk map-based Alzheimer’s disease subtypes. *Note.* **a-c,** Distribution of brain regions with the highest disease risk for the three AD subtypes. The brain anatomical structures with the highest risks are listed in the bottom right of each of the panels. **d,** Patients of the three AD subtypes in a two-dimensional distribution map based on principal component analysis. Solid circles represent patients with high-risk MRI-based AD classifier scores, while hollow circles represent patients with low-risk MRI-based AD classifier scores. AD, Alzheimer’s disease.

To assess the potential of brain subtypes to predict disease progression, we compared the rate of cognitive decline, measured as the annual change in MMSE score (the difference between baseline and the most recent score divided by the number of years), across brain subtypes. We focused on individuals with baseline MCI diagnoses (*n* = 111), as this phase typically shows the most pronounced cognitive deterioration. Results indicate that cognitive decline rates differed significantly among brain subtypes in the MCI group, even after adjusting for sex, age, and education (*F*(2, 105) = 3.63, *p* = 0.030; **Fig. 6b**). Post-hoc comparisons revealed that MCI patients classified as subtype 3 experienced a faster cognitive decline compared to those in subtype 1 (mean difference = 1.46, SE = 0.69, *t*(105) = 2.12, *p* = 0.036) and 2 (mean difference = 1.52, SE = 0.70, *t*(105) = 2.17, *p* = 0.032). In contrast, no significant differences were observed in baseline MMSE scores across the brain subtypes (*F*(2,105) = 1.56, *p* = 0.212; **Fig. 6a**).

**Fig. 6.**
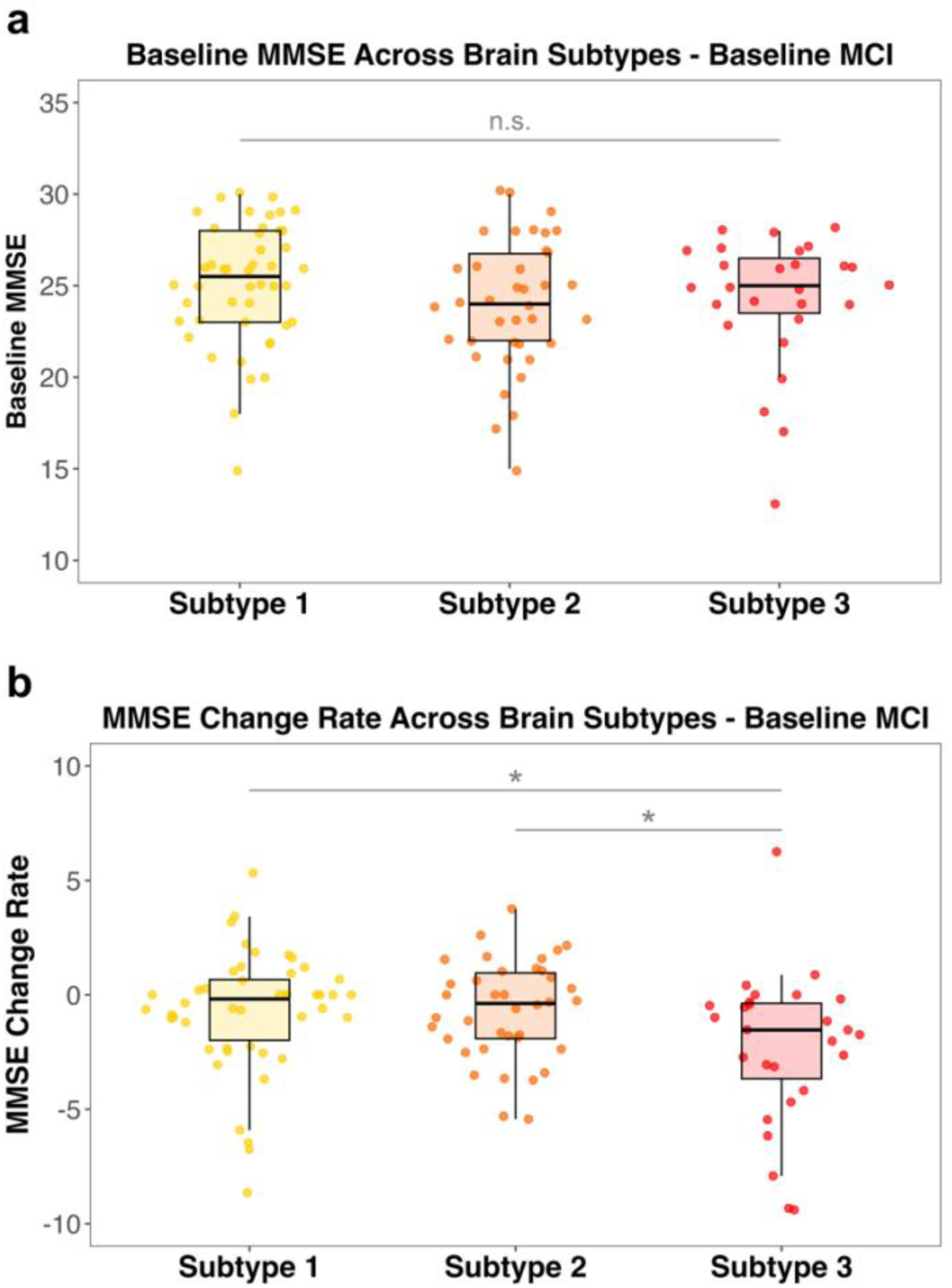
Baseline scores and change rates of MMSE scores across different brain subtype groups in individuals with baseline MCI. *Note*. **a,** Baseline MMSE scores across different brain subtype groups in individuals with baseline MCI; **b,** MMSE change rates across different brain subtype groups in individuals with baseline MCI. MMSE, Mini-Mental State Examination; MCI, mild cognitive impairment. n.s., non-significant, \**p* < 0.05.

Demographics and risk score profiles for different brain subtypes in the MCI group are presented in **Fig. S3**. Cognitive decline rate did not differ significantly among subtypes in the NC or SCD groups (**Fig. S4-5**)

## DISCUSSION

We examined the potential clinical utility of an MRI-based AD deep learning model within a comprehensively phenotyped longitudinal Chinese cohort. The classifier, which had been trained on North American samples, demonstrated good generalizability in distinguishing AD from NC within the Chinese sample, achieving an AUC of 91.3% with high sensitivity.

Importantly, the model accurately identified 86.7% of participants who would progress to AD up to 14 years before their AD diagnosis. MCI patients defined as high risk by the model were significantly more likely to progress to AD. The interpretable model provided personalized brain risk maps and identified the MCI subtype that is most prone to rapid cognitive decline. These predictive abilities enable the use of structural MRI to select patients who are most likely to benefit from early drug intervention. Finally, the MRI-based risk scores were significantly correlated with global cognitive measurements and plasma-based biomarkers.

### AD classification performance and generalizability

The classifier’s ability to accurately distinguish AD from NC in the SILCODE Chinese cohort and previously in four Western samples underscores its robustness and potential for broader application across diverse populations^34^. Currently, MRI is primarily used for staging, monitoring disease progression, and detecting potential adverse events from pharmacological treatments, such as amyloid related imaging abnormalities (ARIA). However, MRI integrated with artificial intelligence is poised to be more closely integrated into screening, early diagnostics and precision interventions for the AD spectrum^45^. Compared to PET scans and cerebrospinal fluid tests, MRI’s non-invasive and radiation-free nature, ready availability, low cost and high sensitivity make it more suitable for large-scale disease screening^46^. This may facilitate the decision-making process for proceeding with monoclonal antibody treatments, thereby significantly advancing the timing and potentially enhancing the effectiveness of interventions. The classifier’s slightly lower performance (AUC of 91.3%) than in the original study is likely due to differences in population characteristics. Age is the most critical factor influencing the model’s predictive performance. The samples used for training and testing the MRI-based models in the previous study (76 ± 7 years in ADNI, 74 ± 7 years in AIBL, 70 ± 7 years in MIRIAD and 75 ± 10 years in OASIS)^17^ were significantly older than the SILCODE samples (66.6 ± 7.6 years). According to the 2024 report by the Alzheimer’s Association, the incidence of AD in the 75-84 age group is 2.64 times higher than in the 65-74 age group^47^. This implies that a considerable number of SILCODE participants with high-risk scores, currently diagnosed as NC, are likely to progress to AD in the future. In fact, this inference aligns with the model’s performance, which demonstrates a remarkable sensitivity of 95.2% but a less robust specificity of 85.6%. These findings underscore the model’s effectiveness in correctly detecting AD, making it well-suited for large scale AD screening and selecting patients for drug therapies. In such scenarios, the priority is to minimize the likelihood of overlooking AD patients and those at high risk for developing AD. Furthermore, considering the classifier can provide an in-advance alert of AD progression risk, it is plausible that at least some of the healthy subjects currently identified as “false positives” might be at a higher risk of developing AD in the future, warranting closer monitoring.

In addition, structural differences in brain gray matter volume and density between Caucasian and East Asian populations^48,49^ may affect classifier performance. Given the limited number of AD patients in SILCODE, we chose not to fine-tune the model so as to avoid overfitting. Nevertheless, testing the model’s generalizability directly in a different population could also be a valuable next step. Furthermore, although the current model was not trained with SCD or MCI patients, the output scores for different clinical diagnoses differed significantly. Higher risk scores corresponded to more advanced clinical stages, highlighting a valuable gradient of AD-related effects.

### Predicting early-stage AD progression by MRI-based risk scores

The results of the survival analysis and lead time analysis further validated that MRI combined with deep learning could effectively predict the progression of early-stage AD patients. Previous research has shown that structural MRI changes can be detected ten years before clinical diagnosis at the group level ^50,51^. In the current study, the classifier was able to detect abnormal scores up to 14 years before diagnosis at the individual level—with 57% of converters showing abnormal scores 5–14 years prior, 78% within 3–5 years, and ∼90% within 3 years. This indicates that the current model can detect early-stage neurodegenerative changes well before the onset of significant cognitive decline and visually observable brain atrophy.

Furthermore, the MCI patients identified as high-risk by the MRI-based risk score showed a significantly higher probability of progressing to AD compared to those classified as low-risk. The median survival time (i.e., time-to-conversion) for the high-risk MCI group (31 months) was significantly shorter than that of the low-risk MCI group (>167 months). This provides a valuable tool to identify high-risk MCI patients who are more likely to benefit from close monitoring and intervention. Emerging treatments, such as those targeting amyloid plaques and tau pathology, are more effective when applied during the earliest phases of neurodegeneration^52,53^. By identifying high-risk individuals early, clinicians can intervene at a stage when disease-modifying therapies may have the greatest impact, potentially delaying AD progression and improving long-term outcomes.

### Subtyping and predicting rate of cognitive decline via an interpretable model

The individualized brain risk maps derived from our interpretable models offer a novel approach to understand and visualize disease pathology and progression at the individual level. The ability to identify specific brain regions affected by AD may facilitate targeted interventions, i.e., personalized medicine, and enhance physician confidence in the deep learning model.

Interestingly, the AD-related risk areas include not only the most commonly affected hippocampus and posterior cingulate cortex, but also encompass many deep structures such as the midbrain tectum, red nucleus, dorsal raphe nucleus, and pineal gland. These brain regions constitute the main risk areas corresponding to Subtype 1. These regions are involved in the processing of audiovisual information and motor control, and regulate circadian rhythms and mood through the secretion of melatonin and serotonin^54–58^. A substantial amount of morphological and pathological evidence shows that these structures are altered in AD, and are closely associated with symptoms such as sleep disturbances, audiovisual integration, and depressed mood. For example, the RNA binding protein, TDP-43, is strongly associated with the morphological and clinical features of AD, and TDP-43 deposited in midbrain tectum is included as a key feature of Stage 5 in the latest AD staging scheme^59,60^. Additionally, the pineal gland in AD patients is significantly smaller than in NC and MCI and its volume is associated with cognitive decline^58^. Furthermore, the subtypes based on the risk map can be used to identify MCI patients with the fastest cognitive decline (Subtype 3). This can provide guidance for selecting MCI patients who are most likely to benefit from pharmacological treatment.

### Physiological and cognitive relevance of the MRI-based risk scores

We also examined the relationship between classifier scores and clinical measurement, including blood biomarkers and cognitive assessments. We used plasma biomarkers from SILCODE due to their greater accessibility in practice than CSF markers. We used the A-T-N-I framework, which stands for Amyloid (Aβ 42/40 ratio), Tau (p-tau 181 and p-tau 217), Neurodegeneration (NfL), and Inflammation (GAFP), to evaluate the physiological relevance of the MRI-based risk score. Among blood biomarkers, we detected significant global correlations between the risk score and the T, N, and I biomarkers. This demonstrates that the classifier scores are not only indicative of structural brain changes observable via MRI but also correspond to biochemical changes occurring in blood.

When analyzing the relationship between risk scores and plasma biomarkers within diagnostic groups, significant associations were observed only in the NC group. This is not surprising, as plasma biomarkers change most rapidly during the preclinical stage^61^. Notably, the association between the risk score and p-tau 181became non-significant when tested within the stages, while the association with p-tau 217 remained significant in the NC stage. This finding aligns with recent studies that identify plasma p-tau 217 as the most accurate and robust biomarker for distinguishing preclinical pathological stages^62^ and predicting the progression of AD among phosphorylated tau markers^63^. Importantly, in the 2024 revision of the NIA-AA criteria, p-tau 217 is the sole plasma biomarker utilized for diagnostic purposes and the only plasma biomarker that is used simultaneously for diagnosis, staging, monitoring progression, and assessing treatment efficacy^20^. The remarkable correlation between the MRI-based risk score and plasma p-tau 217 further validates the diagnostic and screening value of MRI. The significant correlation of MRI-based risk scores with NfL scores is not surprising because brain anatomical structures and NfL levels show concurrent alterations, both manifesting in parallel along the AD progression trajectory^4,5^. The significance of NfL within NC groups aligns with evidence supporting it as a risk biomarker for predicting hippocampal volume changes, a key feature of AD pathology, in cognitively unimpaired individuals^64^.

Interestingly, the amyloid biomarker did not correlate significantly with model scores. This may be because plasma Aβ is not a sufficiently strong biomarker for AD. Supporting this, **Table 1** shows that plasma Aβ42/40 ratios were nearly identical across different diagnoses. Prior studies have similarly reported no significant differences in plasma Aβ42/40 across AD stages and found only weak correlations between plasma Aβ42/40 and its CSF or PET counterparts^65,66^. Differences in methods for measuring plasma Aβ can also impact results. A study comparing various methods and assays found that plasma Aβ42/40 levels measured by the Simoa immunoassay^67^, particularly from Quanterix, as used in the current dataset, do not adequately represent CSF Aβ42/40 or Aβ PET status. Future research should consider using CSF Aβ data or Aβ PET to test the correlation between Aβ and risk scores from our AD classifier, or employ plasma methods that better reflect CSF Aβ levels, such as the immunoprecipitation-coupled mass spectrometry (IP-MS) method^67^.

Although the significant correlations between the MRI-based risk scores and the plasma biomarkers primarily occur in NC samples, the current study cannot entirely rule out the possibility that these associations also occur in MCI population. The small sample size of the MCI individuals with both plasma and MRI data (approximately 30) reduced the likelihood of detecting significant associations.

Associations of MRI-based risk scores with both global cognitive measures (MMSE, CDR, MES, MOCA-B) and subdomain-specific tests (AVLT-L, AVLT-R, AFT, BNT, STT-A, STT-B) were significant, demonstrating the consistency between brain risk and cognitive function.

Within diagnosis, significant associations were observed only during the MCI phase, where cognitive symptoms deteriorate significantly^61^. The remaining significant associations of risk scores with MMSE and MOCA-B, two widely used global cognitive measures in clinical settings, support the clinical applicability of the risk score and suggest that MRI-based risk scores are more closely related to global cognitive status than to specific cognitive subdomains.

### Strength and limitations

The main strength of this study involves the use of a large, longitudinal, well-phenotyped dataset that is independent of the training samples to validate the ability of the MRI-based deep learning model to predict disease progression and cognitive decline. The non-invasive, cost-effective, and widely accessible nature of MRI support expanding its role in AD drug interventions, from safety monitoring for adverse events such as ARIA^68^ to selecting high-priority patients and monitoring drug efficacy. The physiological significance of the MRI-based risk scores is validated by widely accepted and accessible blood-based biomarkers. A few machine learning or deep learning studies have linked MRI-based AD scores with molecular pathology, such as CSF amyloid or tau pathology^69,70^. The current study contributes to this field by providing an evaluation of A-T-N-I pathology in plasma. In the field of neuroregulation, the individualized brain risk maps in this study could help researchers and clinicians pinpoint potential intervention targets. The model has been made open-source and integrated into a free, publicly accessible online platform designed to revolutionize early screening and intervention for AD. By openly sharing our developed models (https://github.com/Chaogan-Yan/BrainImageNet), we empower researchers worldwide to adopt and deploy these tools, fostering global collaboration in AD research. Furthermore, our demonstration website (http://brainimagenet.org) enables users to upload raw T1-weighted MRI scans or preprocessed gray matter density (GMD) and gray matter volume (GMV) data. The platform delivers real-time predictions of AD risk, providing an invaluable resource for clinicians and researchers to identify at-risk individuals early and explore tailored intervention strategies. This initiative underscores our commitment to leveraging advanced technology for proactive, accessible, and scalable AD risk assessment.

Study limitations include the use of a single-site dataset, SILCODE, which hinders effective simultaneous fine-tuning and testing of the model. Future studies should assemble a more extensive and representative multi-center AD dataset to fine-tune the current MRI-based models for better generalizability and to further enhance its value in clinical application. Moreover, the average age of our sample was around 66, which is lower than comparable studies. A future research goal is to build a normative model based on a large sample of healthy individuals to construct age-specific models to improve specificity.

In summary, our MRI-based deep learning AD classifier generalized to a different population, identifying 87% of participants at baseline who progressed to AD. The interpretable models indicated AD risk brain regions and predicted the MCI patients who experienced the fastest cognitive declines. Implementation of MRI in clinical practice can complement current biomarkers such as CSF, PET, and blood tests, facilitating the selection of patients most likely to benefit from prospective monitoring and early immunotherapy interventions, thereby supporting clinical decision-making and precision medicine.

## RESOURCE AVAILABILITY

### Data availability statement

The SILCODE dataset used in this study is not publicly available due to participant privacy and ethical considerations. However, researchers interested in collaboration may contact the corresponding authors to inquire about collaboration opportunities and apply for data access. Applicants are required to provide a detailed research plan and sign a data use agreement. The research team will respond within 30 business days upon receiving applications. All data sharing will strictly adhere to relevant privacy protections and ethical guidelines to ensure the secure and compliant use of the data.

### Code availability statement

The code for the MRI-based deep learning model for Alzheimer’s disease is available at https://github.com/Chaogan-Yan/BrainImageNet. Our demonstration website (http://brainimagenet.org) allows users to upload data directly and obtain risk scores after providing informed consent. Additionally, the codes for statistical analyses, creating individualized AD brain risk maps, and data visualization are available at https://github.com/Chaogan-Yan/BrainImageNet/tree/master/ADPredictionPaperScripts.

## FUNDING AND ACKNOWLEDGEMENTS

Funding for the study was provided by the Sci-Tech Innovation 2030 - Major Projects of Brain Science and Brain-inspired Intelligence Technology (2022ZD0211800 [YH], 2021ZD0200600 [C-GY]), National Natural Science Foundation of China (82122035 [C-GY], 82394434 [YH], 82020108013 [YH], 82327809 [YH]), Scientific Foundation of Institute of Psychology, Chinese Academy of Sciences (E3CX1315 [BL]), Sino-German Cooperation Grant (M-0759 [YH]), Shenzhen Bay Scholars Program (YH), Tianchi Scholars Program (YH), Beijing Nova Program of Science and Technology (20230484465 [C-GY]), Beijing Natural Science Foundation (J230040 [C-GY]), National Institutes of Health grant (U01 AG068057 [P-M-T]).

None of the funding sources had any involvement in the study design, data analysis, or manuscript writing.

## AUTHOR CONTRIBUTIONS

C-GY: Conceptualized and designed the research, interpreted results, revised the manuscript, and supervised the entire research process. YH: Conceptualized the research, interpreted results, acquired the data, and provided expert domain knowledge in the AD research field. JL: Interpreted results, acquired the data, and provided expert domain knowledge in the AD research field. BL: Conceptualized and designed the research, conducted data analysis and visualization, interpreted results, and drafted the manuscript. Y-RC: Designed the evaluation framework, conducted data analysis and visualization, interpreted results, and drafted the manuscript. R-XL, M-KZ, G-QC, and S-ZY: Acquired data and provided diagnostic and data collection details. FXC and PMT: Revised the manuscript.

## Footnotes

1 Model scores derived from linear activation, which are better suited for statistical testing than the default sigmoid-activated risk scores. See Methods for further details.

