## Supplemental File for "Interpretable MRI-Based Deep Learning for Alzheimer’s Risk and Progression"

[Fig. S3. Demographic and risk score profiles across different MCI subtype groups. MCI, mild cognitive impairment. n.s., non-significant, **p < .01, ***p < .001.	3](#_Toc190684122)

Fig. S1. Frequency distribution of intervals between MRI session dates and dates of progression to Alzheimer’s disease (AD) in people who converted to AD during the follow-up period.

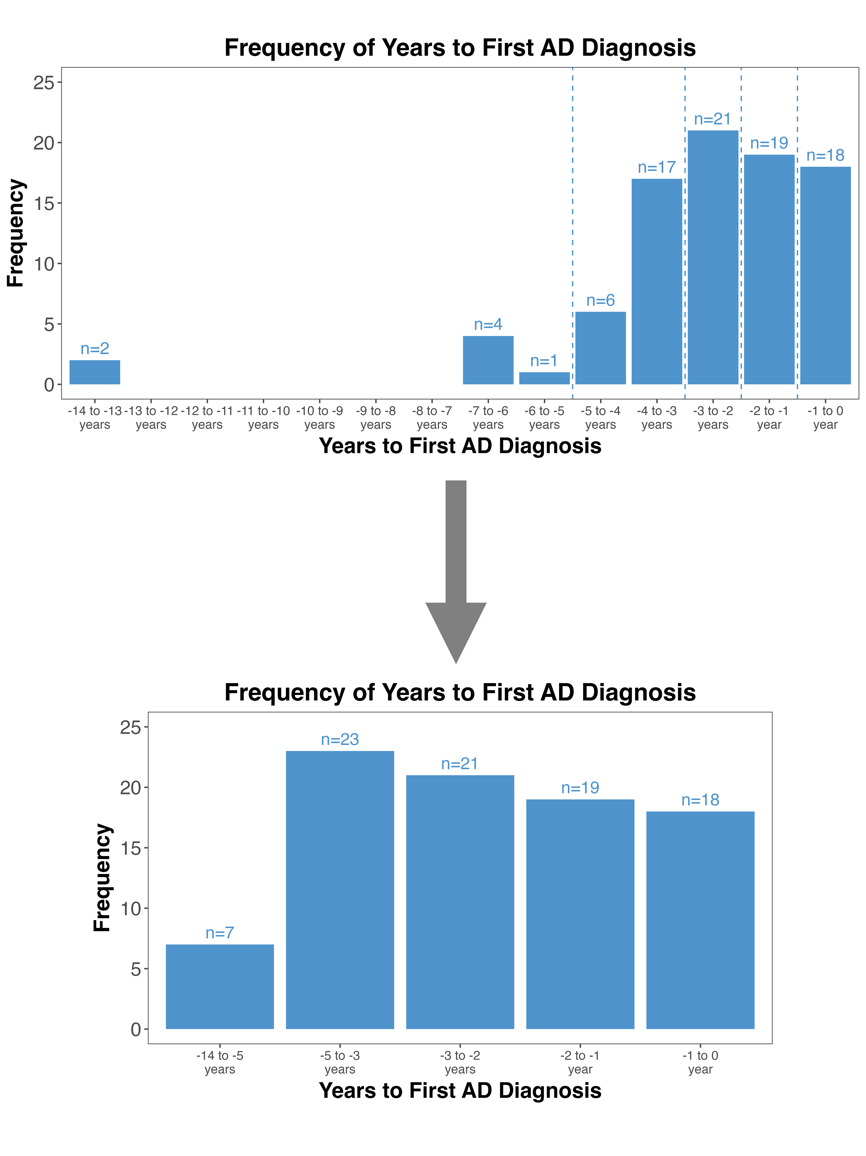

Fig. S2. Sum of Squared Errors at different number of clusters.

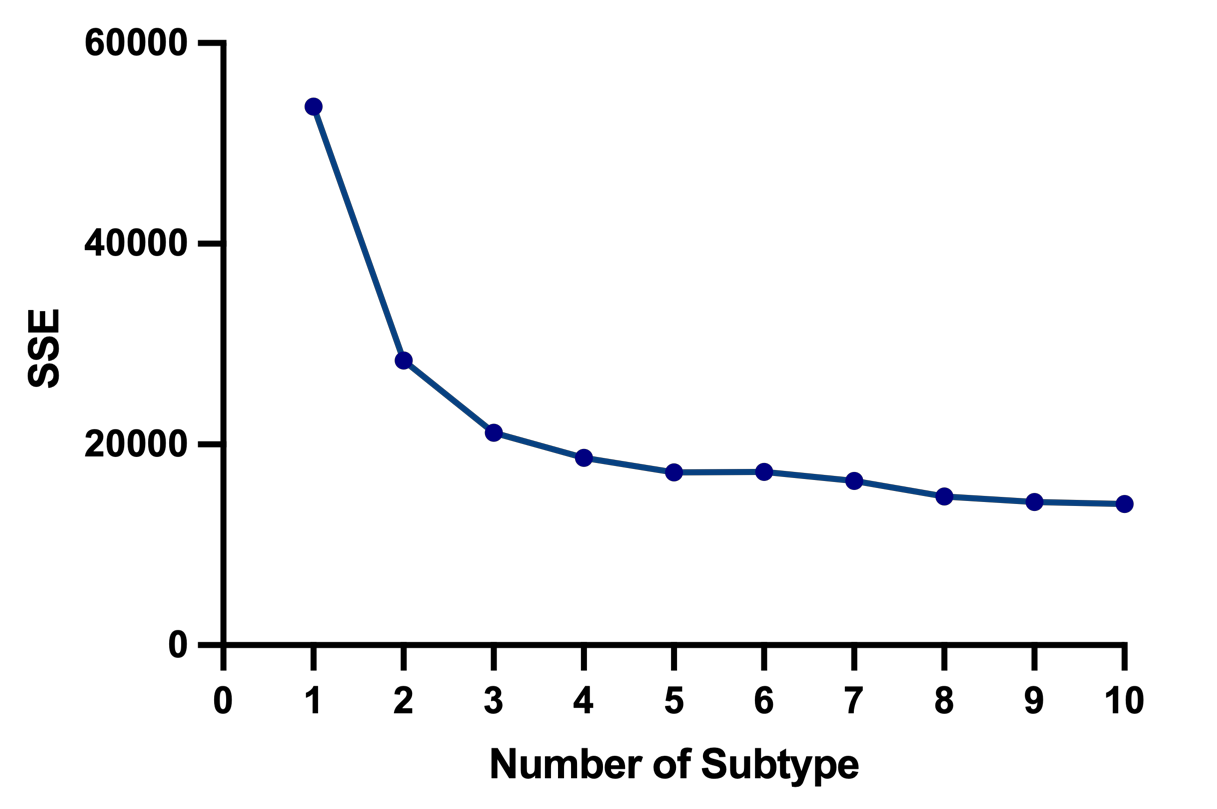

Fig. S3. Demographic and risk score profiles across different MCI subtype groups.

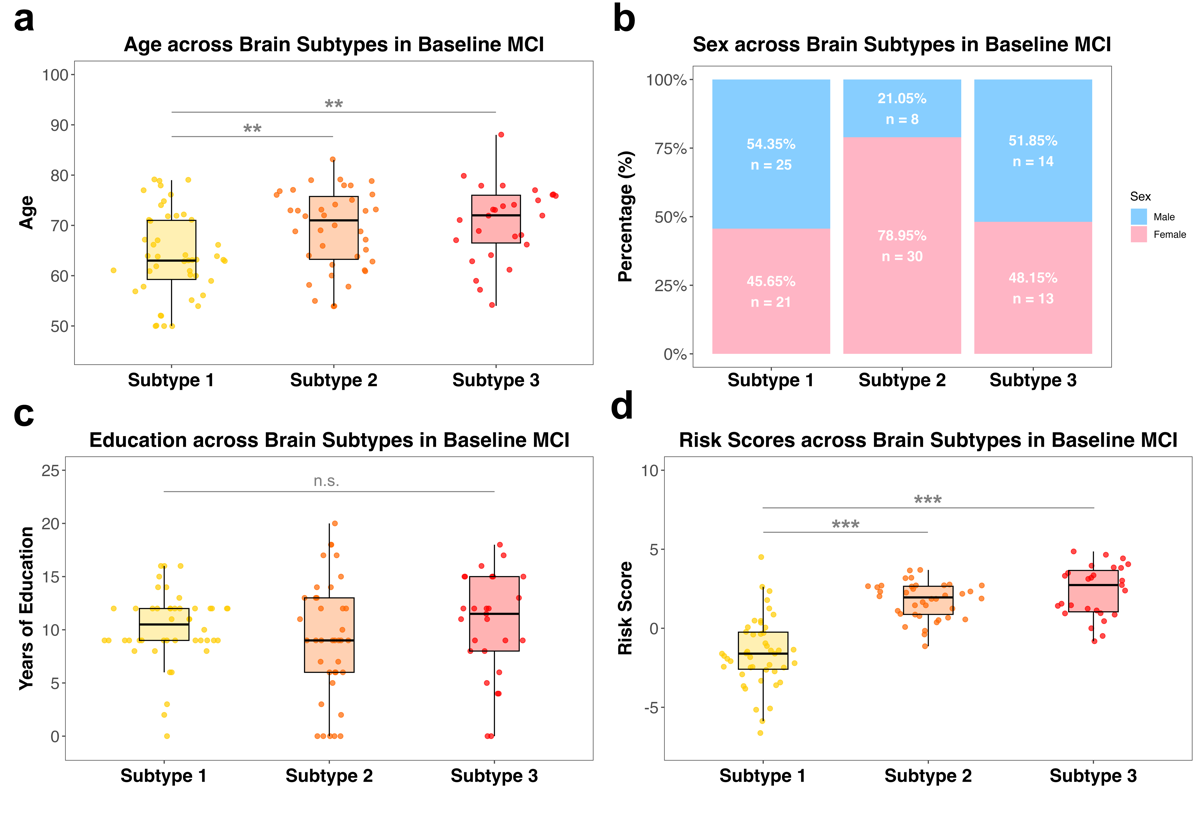

*Note*. MCI, mild cognitive impairment. n.s., non-significant, ***p* < .01, ****p* < .001.

Fig. S4. Baseline scores and change rates of MMSE scores across different NC subtype groups.

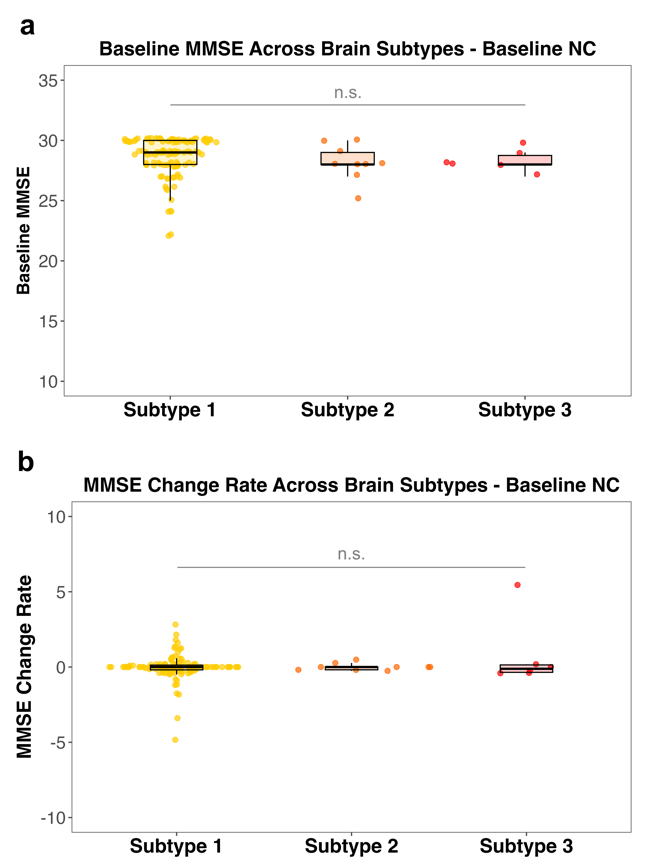

*Note.* NC, normal cognition; MMSE, Mini-Mental State Examination. n.s., non-significant.

Fig. S5. Baseline Scores and Change Rates of MMSE Scores across Different SCD Subtype Groups.

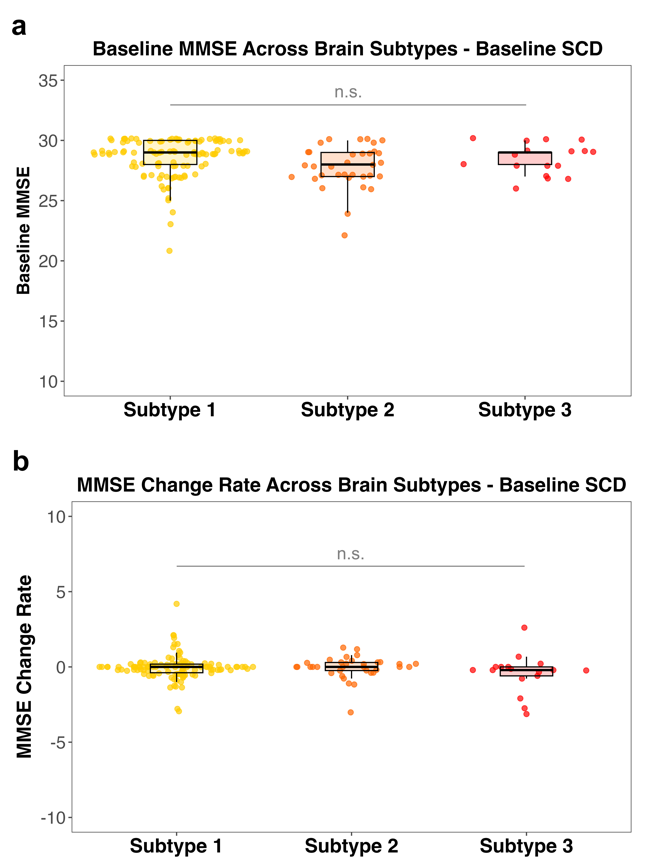

*Note*. SCD, subjective cognitive decline; MMSE, Mini-Mental State Examination. n.s., non-significant.
